# How do inter-organisational electronic health records affect hospital physician and pharmacist decisions? A scoping review

**DOI:** 10.1101/2021.09.09.21254419

**Authors:** Philip Scott, Haythem Nakkas, Paul Roderick

## Abstract

**Objective:** To provide an overview of the effects of inter-organisational electronic health records on inpatient diagnosis and treatment decisions by hospital physicians and pharmacists.

**Materials and Methods:** Five-stage scoping review, using distributed cognition and the information value chain as guiding conceptual models. Eligibility criteria: empirical studies addressing how shared health records were used in inpatient clinical decision-making, published 2008-18. Sources: Healthcare Databases Advanced Search, covering nine sources including PubMed. Charting methods: data extraction form completed by one author, with inter-rater reliability assessment at title and abstract review.

**Results:** Quantitative studies (n=14) often reported relatively low usage of shared records (6.8% to 37.1% of cases). Usage is associated with reduction in diagnostic testing and readmission and variable effects on admissions and overall costs. Qualitative studies (n=6) reported avoidance of duplicate diagnostics, changing clinical decisions, the value of historical laboratory results and optimising the timeliness of care. We found no explicit use of explanatory theoretical models, but there is implicit evidence of an information value chain. We found only one study specifically about pharmacists.

**Discussion:** Relatively low usage is due to clinical judgement whether “extra” data is needed, given current knowledge of the presenting condition and relative complexity. We suggest that extensive EHRs need recommender systems to highlight (sometimes unexpected) relevant content, in parallel with professional guidance on indications for consulting shared records.

**Conclusions:** Clinicians only consult shared health records when they must. Mixed effects on process outcomes are due to the hidden variables of patient complexity, clinician judgement and organisational context.

## BACKGROUND

The adoption of electronic health records (EHRs) is foundational in the shift to “digital health”.^1^ This basic step is necessary to achieve higher-level aspirations such as computerised clinical decision support,^2^ precision medicine^3^ and Learning Health Systems.^4^ Often, electronic health records are limited in scope to a single healthcare provider organisation in a particular care sector, such as hospital, primary care or community care. However, there is also widespread interest in sharing data between primary, secondary, tertiary and social care across large geographical areas to support unscheduled care and services for patients who are treated by multiple healthcare provider organisations.^5, 6^ This is often called “Health Information Exchange” (HIE), which has been variously defined,^7^ but we use the simple definition that it is technology that “allows doctors, nurses, pharmacists, other health care providers and patients to appropriately access and securely share a patient’s vital medical information electronically”.^8^ Unfortunately, it has remained unclear when or how such HIE or an extensive inter-organisational health record is more helpful than local records or a patient summary.^9-11^

The authors were commissioned to conduct an evaluation of a regional inter-organisational health record developed by the NHS in southern England. This shared record, created in the early 2000s, was originally called the Hampshire Health Record^12^ but has since expanded in geographical scope and content so is now called the Care and Health Information Exchange (CHIE).^13^ Regional shared records have fairly recently become national policy for the NHS in England.^5^ One of the specified objectives in the study protocol was “To produce a scoping review of the literature that identifies, appraises and synthesises knowledge about the mechanisms of action of inter-organisational electronic health records on clinical decision-making”. As the scope of our primary research was to evaluate the impact of CHIE on decisions about inpatient diagnosis and treatment, we limited this scoping review to hospital physicians and pharmacists. In this review, the terms “inter-organisational health records”, “shared EHRs” and “Health Information Exchange” (HIE) are generally used interchangeably. This scoping review aimed to inform our primary research by providing an overview of how inter-organisational EHRs can support improvements in direct patient care.

## MATERIALS AND METHODS

We have previously reported our protocol^14^ but summarise the key elements here for convenience. The study followed the five-stage Arksey and O’Malley framework for scoping reviews:^15^ (1) identifying the initial research questions, (2) identifying relevant studies, (3) study selection, (4) charting the data, and (5) collating, summarising and reporting the results. Stages 3-5 are reported in the Results section. We have followed the PRISMA-ScR checklist for reporting scoping reviews.^16^ We adopted sociotechnical systems thinking^17^ and in particular the notion of distributed cognition^18^ as the guiding conceptual models for the review. Specifically, we used the Distributed Cognition for Teamwork – Concentric Layers model (DiCoT-CL)^19^ (**Figure 1**) and the information value chain concept (**Figure 2**).^20^ We particularly wanted to see if there was evidence in the literature relating the sharing of data between organisations with the notion of “team cognition” or with the chain of events from patient interactions to decisions to outcomes.

**Figure 1.** Distributed Cognition for Teamwork – Concentric Layers (adapted from Ref.19 with permission). EHR, electronic health record.

**Figure 2.** Information value chain (reproduced from Ref. 20 with permission).

The ambiguity of terminology mentioned above is one reason why we chose to conduct a scoping review rather than a systematic review. It has been suggested that the indications for choosing the scoping review methodology include clarification of key concepts and definitions in the literature and determining key characteristics or factors related to a concept.^21^ Although we formulated specific research questions (as recommended in the framework and PRISMA-ScR checklist), our aim was fundamentally to provide an overview of knowledge in this field as background for our primary research rather than to synthesise evidence-based guidelines for clinical practice.

A patient and public involvement (PPI) group was set up to advise the development of the project, including this scoping review. The review methods were discussed with the project PPI group and with a regional Young Adults PPI group organised by the South Central Research Design Service of the National Institute for Health Research. These discussions confirmed that the proposed scope was important and relevant to patients and that the approach was satisfactory.

We used the Healthcare Databases Advanced Search (HDAS) resource, provided by the National Institute for Care Excellence and Health Education England, as it enables search of nine relevant databases including PubMed^14^. Articles were included if they were empirical studies or systematic reviews that addressed how inter-organisational electronic health records or health information exchanges were used in inpatient clinical decision-making.

Studies were excluded if they were discussing solely technical aspects or if they addressed only electronic health records within a single organisation. The date range in our search strategy was from April 2008 until April 2018, as both inter-organisational electronic health records and health information exchanges are relatively new innovations.

To chart the results, we developed a data extraction form which was completed by one author, with independent review of a sample of 200 papers at both title and abstract review stages to determine inter-rater reliability.

In stage 1, we identified the research questions. The main research question was: (RQ1) “How do inter-organisational electronic health and care records affect decision-making by hospital physicians and pharmacists?” We also defined secondary research questions: (RQ2) “When are rich electronic health records more useful than summary records?” and (RQ3) “What specific pathways or protocols demonstrate cost reduction or quality improvement (QI) from inter-organisational electronic health records?”

Stage 2 defined the search terms and inclusion/exclusion criteria. We used the following search terms to capture a broad range of relevant literature: ((“Decision-making” OR “Clinical decision-making” OR “Computer-assisted decision-making” OR “clinical decision support systems”) AND (“Medical Records Systems, Computerized” OR “Electronic Health Records” OR “Hospital Information Systems” OR “Health Information Exchange”)). We also hand-searched using the reference lists of the included studies in order to identify additional relevant articles. In addition to our search strategy, we adopted a highly relevant review by Bowden & Coiera as a supplementary source.^22^ We also included other systematic reviews that we found in our search, so that we could compare their scope and conclusions with this review.

## RESULTS

Stage 3 was the search execution. Using the search terms specified in stage 2, 2196 articles were identified from PubMed and 563 from other HDAS sources. Based on title and abstract review, we excluded the majority of these, but a further 14 articles were identified from the Bowden & Coiera review. Search results were downloaded and imported into Microsoft Excel for further analysis. Guided by the inclusion and exclusion criteria, 22 studies were identified as being pertinent to the review. Two extra snowball references were located. Full text versions of the articles were obtained, with each article being reviewed and confirmed as appropriate by all authors. In total, 24 studies were included in the review. **Figure 3** below shows the PRISMA diagram.

**Figure 3.** PRISMA diagram for study selection.

The inter-rater reliability assessment at title and abstract review stage, based in each case on a sample of 200 papers, resulted in Cohen’s kappa = 0.71 showing substantial agreement.^23^We re-assessed the papers where we had variant conclusions and agreed on all final inclusion and exclusion decisions. At the full text review stage, we jointly assessed all papers and so did not calculate inter-rater reliability.

In stage 4 we charted the data. This search strategy yielded 3 systematic reviews and 21 articles from five countries. Thirteen of the studies were primarily quantitative, 6 were primarily qualitative and one used mixed methods. The quantitative studies were from the USA (n=9) and Israel (n=6). The qualitative studies were from the USA (n=4), UK (n=2) and Canada (n=1).

**Table 1** summarises the findings of the 3 systematic reviews. **Table 2** presents the results of the 14 quantitative studies and the quantitative results of the mixed methods study. **Table 3** gives the key conclusions of the 6 qualitative studies and the qualitative results of the mixed methods study.

**Table 1.** Key findings of the systematic reviews.

| Ref | Authors | Scope | Date range | N studies included | Findings |
| --- | --- | --- | --- | --- | --- |
| 24 | Rudin et al., 2014 | Use and effect of HIE on clinical care. | 2003-2014 | 12 hypothesis-testing studies<br>12 studies of HIE usage<br>17 studies of financial sustainability<br>38 studies of attitudes and barriers | Low-quality evidence of cost savings in ED<br>Wide variation in usage levels: typically, 2-10%, but one site achieved 60%<br>25% claim to have sustainable business model<br>HIE considered valuable but barriers remain: costs, privacy concerns, technical and workflow issues |
| 22 | Bowden & Coiera, 2017 | Impact on accessing primary care records in unscheduled care. | No limit | 22 | Shared EHRs poorly evaluated. Heterogeneity of systems and populations make it difficult to generalise. No studies used any theoretical model. Evidence for shared EHR benefits is weak. |
| 25 | Hersh et al., 2015. | HIE outcomes. | 1990-2015 | 34 | No studies reported on clinical outcomes or identified harms. Low-quality evidence generally finds that HIE reduces duplicate laboratory and radiology testing, emergency department costs, hospital admissions and improves public health reporting, ambulatory quality of care and disability claims processing. Most clinicians attributed positive changes in care coordination, communication and knowledge about patients to HIE. |

**Table 2.**
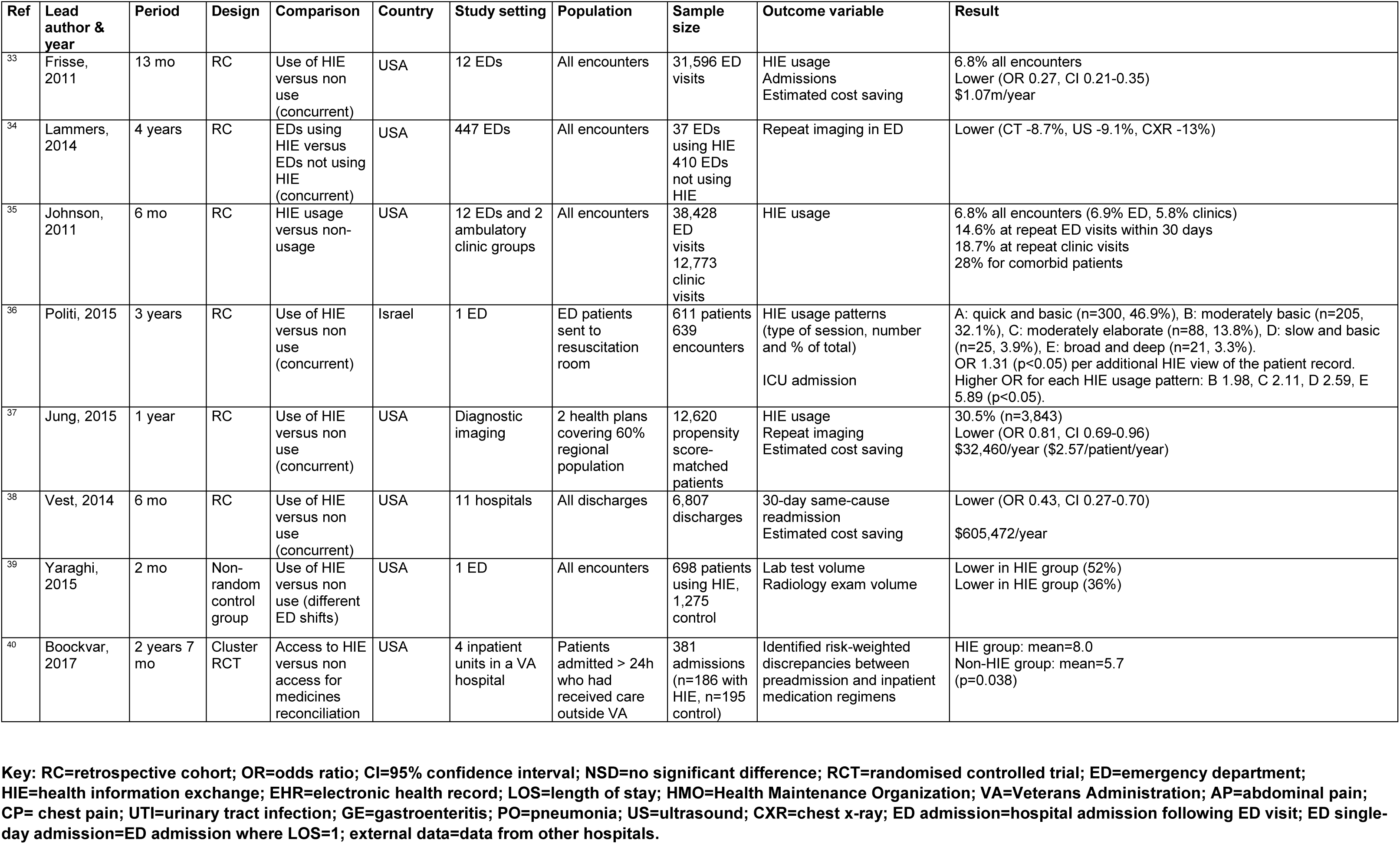
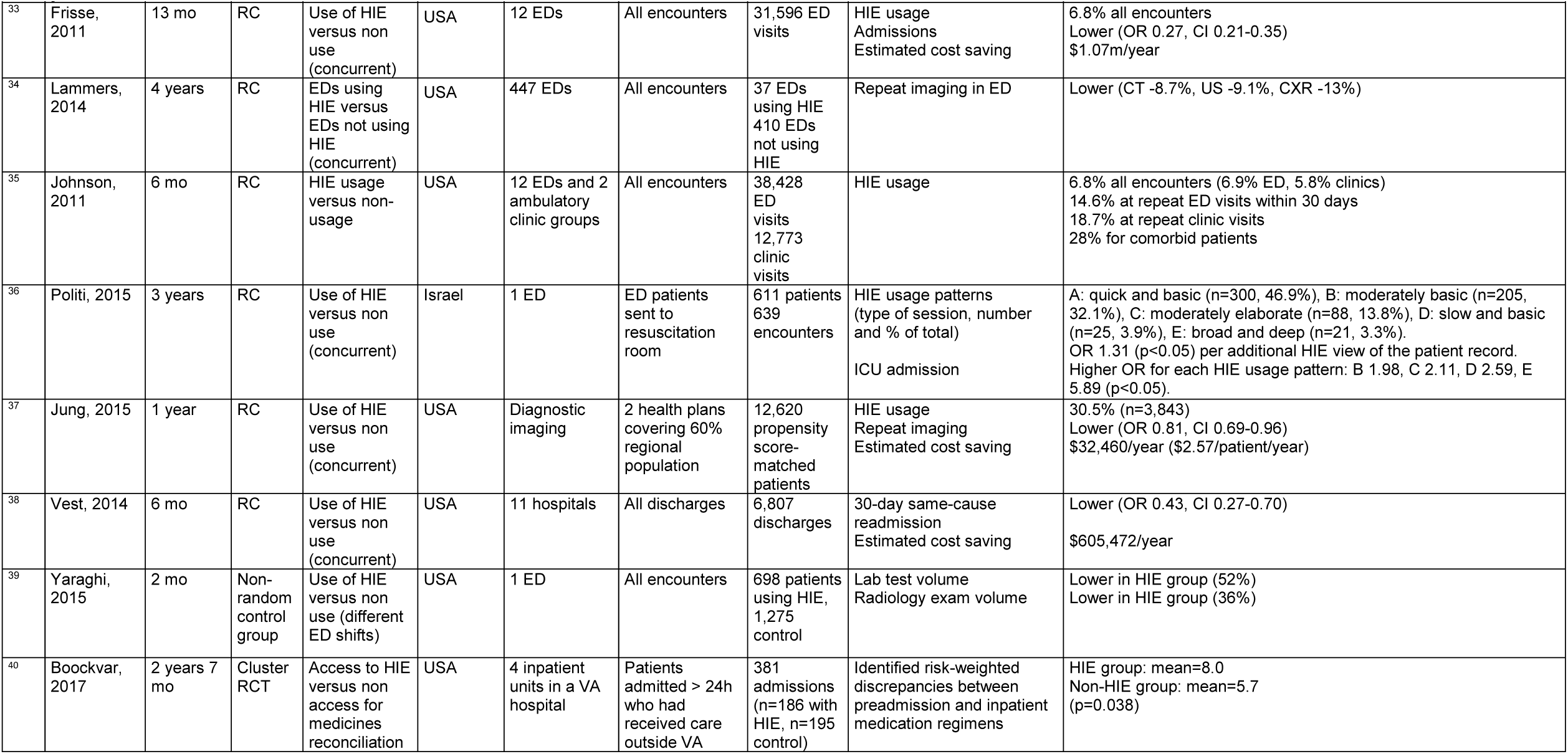
Key quantitative results.

**Table 3.**
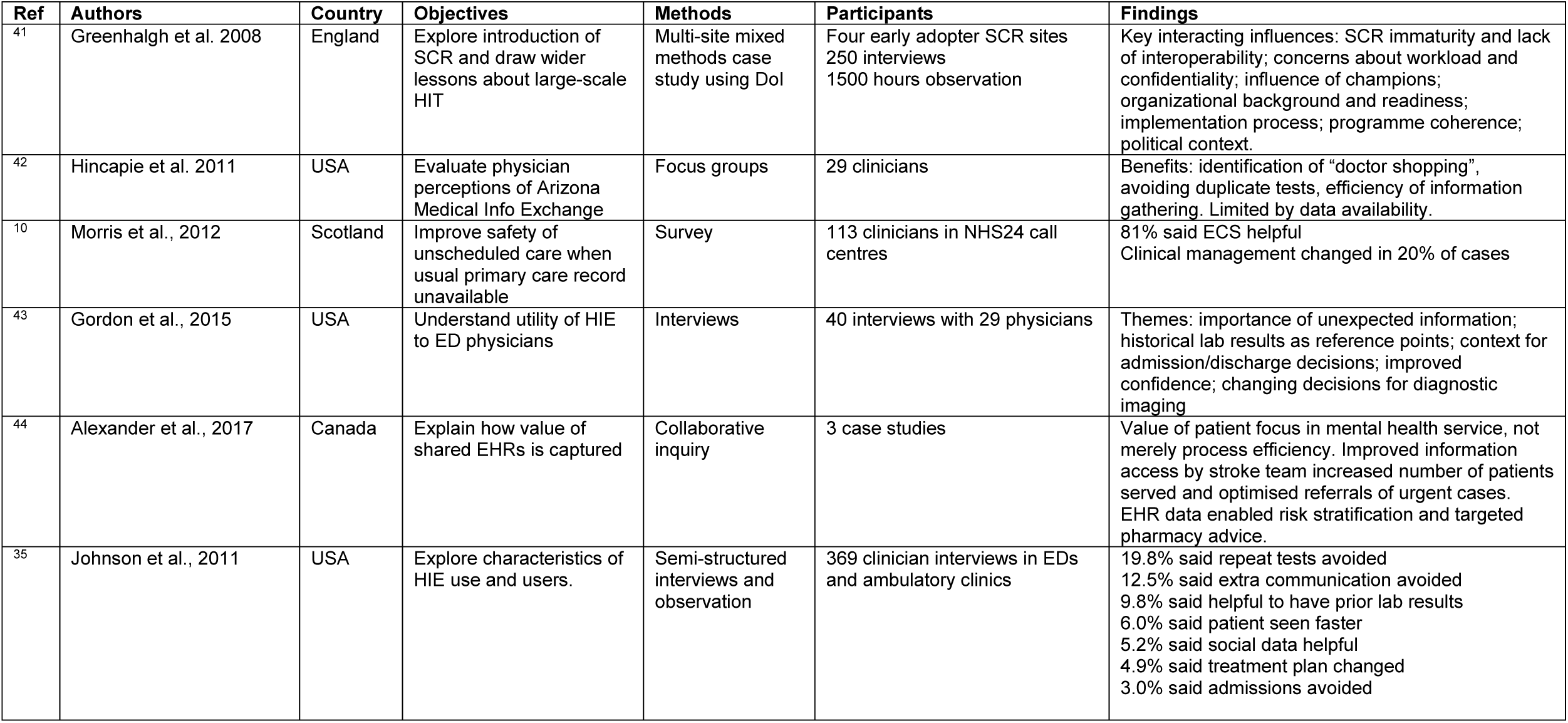
Key qualitative results.

| Ref | Authors | Country | Objectives | Methods | Participants | Findings |
| --- | --- | --- | --- | --- | --- | --- |
| <sup>41</sup> | Greenhalgh et al. 2008 | England | Explore introduction of SCR and draw wider lessons about large-scale HIT | Multi-site mixed methods case study using DoI | Four early adopter SCR sites<br>250 interviews<br>1500 hours observation | Key interacting influences: SCR immaturity and lack of interoperability; concerns about workload and confidentiality; influence of champions; organizational background and readiness; implementation process; programme coherence; political context. |
| <sup>42</sup> | Hincapie et al. 2011 | USA | Evaluate physician perceptions of Arizona Medical Info Exchange | Focus groups | 29 clinicians | Benefits: identification of “doctor shopping”, avoiding duplicate tests, efficiency of information gathering. Limited by data availability. |
| <sup>10</sup> | Morris et al., 2012 | Scotland | Improve safety of unscheduled care when usual primary care record unavailable | Survey | 113 clinicians in NHS24 call centres | 81% said ECS helpful<br>Clinical management changed in 20% of cases |
| <sup>43</sup> | Gordon et al., 2015 | USA | Understand utility of HIE to ED physicians | Interviews | 40 interviews with 29 physicians | Themes: importance of unexpected information; historical lab results as reference points; context for admission/discharge decisions; improved confidence; changing decisions for diagnostic imaging |
| <sup>44</sup> | Alexander et al., 2017 | Canada | Explain how value of shared EHRs is captured | Collaborative inquiry | 3 case studies | Value of patient focus in mental health service, not merely process efficiency. Improved information access by stroke team increased number of patients served and optimised referrals of urgent cases. EHR data enabled risk stratification and targeted pharmacy advice. |
| <sup>35</sup> | Johnson et al., 2011 | USA | Explore characteristics of HIE use and users. | Semi-structured interviews and observation | 369 clinician interviews in EDs and ambulatory clinics | 19.8% said repeat tests avoided<br>12.5% said extra communication avoided<br>9.8% said helpful to have prior lab results<br>6.0% said patient seen faster<br>5.2% said social data helpful<br>4.9% said treatment plan changed<br>3.0% said admissions avoided |

Stage 5: Collating, summarising and reporting the results. We found six recurring themes related to our research questions from the quantitative results given in **Table 2**. We define themes as similar findings in three or more studies. The derived themes are shown in **Table 4**. Overall, low usage was the most frequently reported finding, with a minimum of 6.8% of encounters, a maximum of 37.1% and mean 20.8%. This is relevant as it relates to the prior decision about when HIE usage is required. There were superficially contradictory effects on admission decisions. Each of these six themes is at least implicitly related to some impact of inter-organisational health records upon clinical decision-making.

**Table 4.**
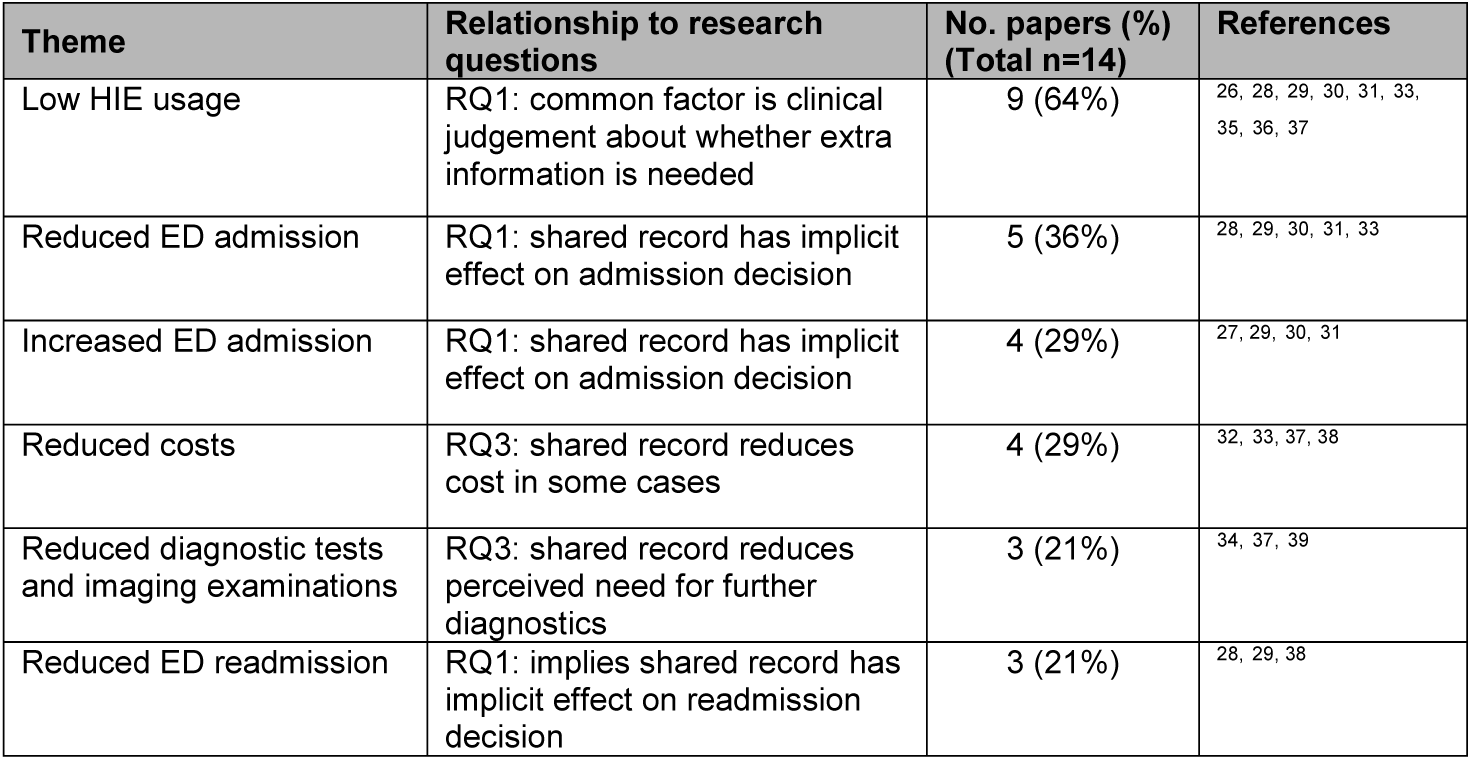
– Recurring themes in the 14 quantitative studies.

Other interesting findings that only featured in one or two quantitative studies, but relevant to our research questions, were: improved guideline adherence,^26^ reduced ED length of stay,^32^ increased detection of medication discrepancies,^40^ costs unchanged,^26 32^ variation in the EHR components used according to differential diagnosis^30^ and the existence of distinct patterns of usage (ranging from “quick and basic” to “broad and deep”).^36^

These findings are consistent with four qualitative findings: avoiding duplicate diagnostics,^35, 42^ changing clinical decisions,^10,35^,^43^ the value of historical laboratory results,^35,43^ and optimising the timeliness of care.^35,44^

## DISCUSSION

### RQ1: How do inter-organisational electronic health and care records affect decision-making by hospital physicians and pharmacists?

The mixture of positive and negative results about admission rates from ED, ED length-of-stay and diagnostic volumes illustrate the multifaceted interplay of factors at work here. Variance is inferred to arise from the collective impact of richer knowledge about each patient in a particular population. The various net financial impacts reported are consequences of that set of effects. It is not a simple linear relationship of “more data reduces admissions and length-of-stay and therefore cost”. As noted in our protocol, this is a complex socio-technical phenomenon. However, in this scoping review, we found only passing references to socio-technical impacts where the theoretical lens of the DiCoT-CL model^19^ would be directly applicable. This probably reflects more about how studies were framed than the actual causal factors at work, given that many benefits described as linear effects in fact require multiple team interactions (for example, avoiding readmission or reducing length-of-stay). Bowden & Coiera^22^ also noted the absence of theoretical framing in the evaluation of inter-organisational health records. One qualitative study^41^ did take an explicitly socio-technical perspective, but concentrated on factors related to adoption rather than effects on clinical decisions.

One qualitative study^43^ specifically explored the impact of HIE upon clinical decision-making in the emergency department. Clinicians reported that using HIE changed their decision-making in 12/37 (32%) of the patient encounters studied. Notably, in 34/37 (92%) of the encounters, the clinician was looking for a specific “known unknown”, but in 14/37 (38%) cases found unanticipated useful information. Clinicians reported increased confidence, even when a decision was unchanged, and that more complete information gave context to decisions. In comparison, another qualitative study reported changed clinical decisions in 4.9% of cases^35^ and yet another reported 20%.^10^

A cluster of findings suggests that the generally observed low rate of usage of inter-organisational health records is predominantly down to clinical judgement about whether extra information is needed for a particular patient. The literature showed usage variance by differential diagnosis^30^, usage patterns ranging from “quick and basic” to “broad and deep”,^36^ admission rates co-varying with level of ED crowding,^27^ association between admission rates and information component usage^29^ and searches for specific missing data.^43^ In other words, we propose that the low usage rate is largely due to the judgement by the clinician about relative need for more data given what is already known and the nature of the immediate presenting problem. The “hidden variables” in usage levels for inter-organisational health records seem to be patient complexity, clinician judgement and organisational context.

Therefore, there is implicit evidence that an information value chain^20^ exists with respect to inter-organisational health records, in that decisions clearly are changed, processes are altered and outcomes are changed. It is quite obvious, for example, that discovering relevant laboratory test results or imaging examinations simply obviates the need for their repetition. However, the steps in most instances of that chain remain fairly opaque. We have a glimmer of insight from the study which analysed EHR component usage,^29^ which showed a relative increase in admissions for emergency patients with chest pain when admission history and blood pressure were viewed and a relative reduction when laboratory results and imaging reports were viewed. It does seem plausible that there would be an association between presenting condition and perceived information need. What we do not know is how each clinician decided which parts of the record were important for which chest pain patients and whether such judgements are consistent professional practice across a clinical team or merely idiosyncratic. This current limited understanding of the information value chain is tentative and solely qualitative, given the heterogeneity of the quantitative results.

This review found limited data about the impact of inter-organisational health records on pharmacist decisions. We know anecdotally that shared records are frequently used in hospital medicines reconciliation, but we found only one published study of this aspect. There was one other reference to medication, in a study that mentioned shared EHR data enabling risk stratification and targeted pharmacy advice.^44^

### RQ2 When are rich electronic health records more useful than summary records?

We cannot answer this question precisely from the literature, but there are some straightforward inferences that can be drawn. Firstly, the overall low usage rate of inter-organisational health records in itself implies that only the minimum necessary set of data is usually wanted. This is supported by the finding^36^ that the “quick and basic” use pattern of HIE and the “moderately basic” use pattern between them account for 79% of usage and the observation^43^ that in 92% of patient encounters, the clinician was looking for a specific item of missing data. Related earlier work has suggested a very limited “lifetime” of information utility.^16^ Secondly, there are some examples of when more than basic data is needed: extant history of previous admissions or surgery^29^ or when presenting complaint is chest pain, abdominal pain or gastroenteritis.^28^ A prospective evaluation by clinical scenario and regression analysis of usage levels and patient parameters would be needed to make any robust conclusions about this question, but there is an obvious relationship between the complexity of the case and the perceived relative need for rich data. However, there is also the finding^43^ that in 38% of cases the clinician found information that was *unexpectedly* useful, so there is also a need to signpost potentially relevant data once there are some decision rules for how to work out what that is.

### RQ3 What specific pathways or protocols demonstrate cost reduction or quality improvement from inter-organisational electronic health records?

We found no consistent evidence of particular pathways or protocols where costs were reduced or quality was improved by using inter-organisational electronic health records. There is consistent evidence of reduction in laboratory tests and diagnostic imaging, but the only specific condition where that was shown is adult repeat emergency visit after headache.^26^

### Comparison with previous reviews

All three systematic reviews that we considered highlighted the relative weakness of evidence about this topic. The substantial variation in HIE usage levels has been noted,^24^ as has the absence of theoretical framing^22^ and data about patient outcomes.^25^ Two out of three systematic reviews noted that clinicians were generally positive about the value of shared EHRs.^24,25^ Two of the systematic reviews mentioned the role of HIE in reducing medication discrepancies.^22,25^

### Further work

We suggest that future studies of shared EHRs and clinical decision-making should explicitly address the socio-technical concept of team cognition, as modelled in DiCoT-CL, and the related notion of “collective intelligence”.^45^

We hypothesise that it is feasible to estimate a maximum achievable usage level of inter-organisational health records by developing regression models of actual usage and population case-mix. There is the potential to use propensity scoring^46^ to model patient populations to control for “confounding by indication”,^47^ using covariates such as presenting complaint, polypharmacy, comorbidity, surgical history and last admission.

Similarly, we propose that should be possible to develop decision algorithms for information that is potentially useful in a given case but may not be obvious to the clinician from their immediate knowledge. There are significant data analysis and usability design challenges in somehow highlighting the additional unexpected information, but this feature would facilitate targeted quality improvement work that aimed to change clinical judgements about when to use a shared record. This would, in effect, be a form of “recommender system”.^23^ This could be developed in parallel with professional guidance on indications for consulting shared records, with exemplars of the type of information that can change practice. There should also be consideration of how to bring this kind of awareness into healthcare professional training.

Finally, we suggest a new set of research questions: How many more patients could have reduced diagnostics if inter-organisational health records were used routinely? What is the cost/benefit trade-off of time to find the “extra” data versus cost avoidance and reduced treatment burden? What are the patient safety benefits especially for people who are confused or unconscious?

### Limitations

There is a notable concentration of literature from only a small number of developed countries, presumably reflecting both the nature of their healthcare systems and the relative levels of investment in regional information sharing. As this is a scoping review, we did not make a quality appraisal of the included studies. Our scope included hospital pharmacists, but we found only one study featuring this profession.

We found no follow-up information of longer-term impact on patient clinical outcomes or quality of life. All the studies looked at process outcomes. The study designs were all observational, mostly retrospective cohorts with varying comparisons (mostly concurrent or before-after) and varying adjustments for confounding between patients. There is limited data on what was accessed and how it was used to change decisions.

We note that the specific issues around inter-organisational EHRs may not be intrinsically different from mono-organisational EHRs that contain many years of patient data. There are perhaps questions of provenance and trust and variations in the quality and consistency of both structured and unstructured data, but we did not explore these in detail.

## CONCLUSIONS

In the literature that we reviewed, clinicians did not use inter-organisational electronic health records routinely but only when they judged it really necessary. There are mixed effects on admission and costs partly due to confounding by indication and the hidden variables of patient complexity, clinician judgement and organisational context. Health IT programmes should be realistic about what a shared health record can and cannot achieve, what constitutes a satisfactory level of usage and seek to gather data on patient outcomes.

## Data Availability

The data that support the findings of this study are available from the corresponding author, [PS], upon reasonable request.

